# Dyslipidemia and development of chronic kidney disease in non-diabetic Japanese adults: a 26-year, community-based, longitudinal study

**DOI:** 10.1101/2024.10.25.24315968

**Authors:** Yukari Okawa, Toshiharu Mitsuhashi

## Abstract

Follow-up studies evaluating the relationship between dyslipidemia and chronic kidney disease (CKD) in non-diabetic populations are limited. This longitudinal study (1998–2024) examined whether the prevalence of dyslipidemia is associated with the subsequent development of CKD in non-diabetic Japanese adult citizens of Zentsuji, Kagawa Prefecture, Japan. Dyslipidemia was defined as low-density lipoprotein cholesterol concentrations ≥140 mg/dL, high-density lipoprotein cholesterol concentrations <40 mg/dL, and/or triglyceride concentrations ≥150 mg/dL. Participants were considered to have developed CKD if their estimated glomerular filtration rate was <60 mL/min/1.73 m^2^. The proportional hazard assumptions were violated. Therefore, the Weibull accelerated failure-time model was selected using the Akaike and Bayesian information criteria. The final cohort included 5970 participants, 41.6% of whom were men. The mean follow-up was 7.09 years. After the follow-up, 1890 (31.7%) participants developed CKD. Participants with dyslipidemia had a 5% shorter survival time (95% confidence interval: 3%–7%) to incident CKD compared with those without dyslipidemia in the full model. High-density lipoprotein cholesterol concentrations <40 mg/dL and triglyceride concentrations ≥150 mg/dL also reduced the survival time to CKD onset by 5%–6%. Our results indicate that controlling the lipid profile to an appropriate range may contribute to reducing the risk of future onset of CKD.

## Introduction

Dyslipidemia is often comorbid with chronic kidney disease (CKD)[1,2]. However, the causal relationship between dyslipidemia and incident CKD remains controversial. Follow-up studies from the United Kingdom (UK) Biobank (n = 373,523) and China (n = 5183) reported that dyslipidemia was a risk factor for subsequently developing CKD[3,4]. In contrast, in a cohort study of the Japanese general population (n = 289,462), the exposure and outcome variables were reversed; prevalent CKD was a risk factor for high triglyceride (TG) concentrations and low high-density lipoprotein cholesterol (HDL-C) concentrations[5].

One explanation for this disagreement between studies may be different definitions of CKD used in each study. An estimated glomerular filtration rate (eGFR) <60 mL/min/1.73 m^2^ and CKD diagnosis based on health records were used in the UK Biobank study, an eGFR <60 mL/min/1.73 m^2^ was used in the Chinese study, and an eGFR <60 mL/min/1.73m^2^ and/or prevalent proteinuria was used in the Japanese study[3–5]. To assess whether dyslipidemia or CKD occurs first, further evidence from longitudinal studies is required.

Another factor that may affect this consistency between studies is diabetes. Insulin deficiency and hyperglycemia due to diabetes cause changes in plasma lipoproteins, leading to dyslipidemia, and hyperglycemia due to diabetes constantly damages renal vessels, causing CKD[6,7]. Moreover, prevalent diabetes is associated with various diseases (e.g., diabetic nephropathy, cancer, and death), which cause bias when investigating the relationship between dyslipidemia and CKD[8]. Therefore, removing the effect of diabetes by restricting participants to those without diabetes is effective in examining the relationship between dyslipidemia and incident CKD in more detail, rather than simply adjusting for diabetes as a covariate.

In recent years, the global prevalence of dyslipidemia and CKD has been high, with estimated percentages of 34.1% and 9.1%, respectively, which vary by race[9,10]. Additionally, prevalent CKD accounted for 1.2 million deaths worldwide in 2017[10]. Therefore, determining the relationship between dyslipidemia and CKD by race for the primary prevention of CKD and health promotion is important. This longitudinal analysis aimed to examine the relationship between dyslipidemia and new-onset CKD in a non-diabetic adult Asian population.

## Materials and Methods

### Participants

This population-based, longitudinal study used data from an annual checkup (1998–2024) of Zentsuji City, Kagawa Prefecture, Japan[11]. All citizens aged ≥40 years can have checkups (participation ratio: 30%–40%). These checkups include measurement of height, weight, and blood pressure, urinalysis, a blood test, an electrocardiogram, and a questionnaire about their lifestyle, such as alcohol intake and the smoking status. In Zentsuji City’s pilot initiative to promote the health of younger citizens, this city included citizens aged 35–39 years in the eligible criteria for the 1998–1999 fiscal year only. The checkup followed the protocol by the Ministry of Health, Labour and Welfare of Japan[12]. We extracted the data on 1 July 2024, using the same database as that used in previous studies[13–15].

On the basis of our study aim, the following exclusion criteria were applied: a) non-Japanese citizenship, b) missing data of the prevalence of CKD, c) participants with CKD at study entry, d) missing data of the lipid profile, d) missing hemoglobin A1C (HbA1c) values, e) participants with HbA1c values ≥6.5% at study entry, and f) missing data with a small proportion of missing values (only 3 time points for BMI and 2 for liver enzymes), and g) participants with data at a single time point[16]. Participants who had HbA1c values ≥6.5% during the follow-up were treated as censored[16].

### Variables

#### Exposure variable

The exposure variable was dyslipidemia. The definitions of dyslipidemia vary according to the guidelines[17]. This study included only Japanese participants. Japanese adults tended to have higher low-density lipoprotein cholesterol (LDL-C), HDL-C, and TG concentrations than non-Hispanic Whites[18]. Therefore, to reduce the influence of race in classifying dyslipidemia in the Japanese population, the following Japan Atherosclerosis Society guideline definition of dyslipidemia was selected: LDL-C concentrations ≥140 mg/dL, HDL-C concentrations <40 mg/dL, and/or TG concentrations ≥150 mg/dL[19].

#### Outcome variable

The primary outcome measure was the onset of CKD. CKD was defined as an eGFR <60 mL/min/1.73 m^2^[20]. The three-variable equation for Japanese was used to calculate the eGFR: eGFR (mL/min/1.73 m^2^) = 194 × serum creatinine^-1.094^ (mg/dL) × age^-0.287^ (years) (× 0.739 if female)[16,21]. Serum creatinine concentrations were measured using enzymatic methods.

#### Covariates

To minimize the effect of confounding factors, the following covariates were adjusted for: biological sex (male[reference]/female), age (continuous, in years), overweight or obesity (no[reference]/yes), self-reported alcohol intake (non-drinker or seldom drinker[reference]/drinker), self-reported smoking status (non- or ex-smoker[reference]/smoker), hypertension [no[reference]/yes], HbA1c (continuous, in %), liver function test results (aspartate aminotransferase [AST], alanine aminotransferase [ALT], and gamma-glutamyl transferase [GGT]; continuous, in U/L), and residence area in Zentsuji City (east[reference]/south/Tatsukawa/Fudeoka/central/west/Yoshiwara/Yogita)[19,20,22]. Overweight or obesity was defined as a body mass index (BMI) ≥25 kg/m^2^[23]. The participants were treated as hypertensive if their systolic blood pressure was ≥130 mmHg and/or diastolic blood pressure was ≥80 mmHg[24]. HbA1c values according to the Japan Diabetes Society (JDS) values were converted to HbA1c values according to the National Glycohemoglobin Standardization Program (NGSP) from 1998 to 2012: HbA1c_NGSP_ (%) = 1.02 × HbA1c_JDS_ (%) + 0.25 (%)[25].

### Statistical analysis

Person-years were calculated from the first to the last observation date, or to the date of developing CKD or diabetes. The participants’ characteristics were compared according to prevalent dyslipidemia. Categorical variables were compared by the total number of failures, total person-years, and incidence rate per 1000 person-years. Continuous variables are shown as the mean and standard deviation (SD). Kaplan–Meier curves are stratified by prevalent dyslipidemia.

The parametric model was selected in this analysis because the proportional hazards assumption was violated using the Schoenfeld residuals and the log–log plot[26]. The Weibull accelerated failure-time model was selected under the Akaike and Bayesian information criteria[27,28]. To evaluate the effect of covariates, four adjusted models were applied. Model 1 was adjusted for sex and age. Model 2 was further adjusted for overweight or obesity, self-reported alcohol intake, and self-reported smoking status. Model 3 was further adjusted for hypertension, HbA1c, AST, ALT, and GGT. Model 4 was further adjusted for the residential area of the city. To stabilize the models, a multiplicative term was added if the exposure variable had an interaction effect with sex, age, overweight or obesity, self-reported alcohol intake, self-reported smoking status, hypertension, HbA1c, AST, ALT, or GGT.

Two sensitivity analyses were conducted in this study. First, LDL-C, HDL-C, and TGs were separately treated as exposure variables. LDL-C, HDL-C, and TG concentrations were classified into the following three categories: LDL-C concentrations <120 (reference), 120–<140, and ≥140 mg/dL; HDL-C concentrations <40, 40–<60, and ≥60 mg/dL (reference); and TG concentrations <100 (reference), 100–<150, and ≥150 mg/dL[19]. Second, participants with an eGFR <60 mL/min/1.73 m^2^ and/or prevalent proteinuria ≥1+ were regarded as having prevalent CKD[29].

Missing values of the self-reported smoking status (24.6% of the total observations), self-reported alcohol intake (25.4% of the total observations), and the residential area (1.50% of the total observations) were imputed using multiple imputation methods (30 imputations)[30,31]. In this analysis, a two-tailed p value <0.05 was regarded as statistically significant. All statistical analysis was conducted using Stata/MP 16.1 (StataCorp, College Station, TX, USA). A flowchart of the participants was created by Python 3.11.10. The study followed the Strengthening the Reporting of Observational Studies in Epidemiology reporting guideline[32].

## Results

### Main analysis

This number of initial participants was 15,788 (41.0% men; 34–103 years old; 124,156 observations). After applying the exclusion criteria, our final cohort included 5970 participants (41.6% men; 39–103 years old; 40,942 observations), with a mean follow-up of 7.09 years (**Figure 1** ). Kaplan–Meier curves by the dyslipidemia category are shown in **Figure 2** . The Kaplan–Meier curve showed that the survival rate was slightly lower in the dyslipidemia group than in the normal lipid profile group (log-rank p <0.001).

**Figure 1.**
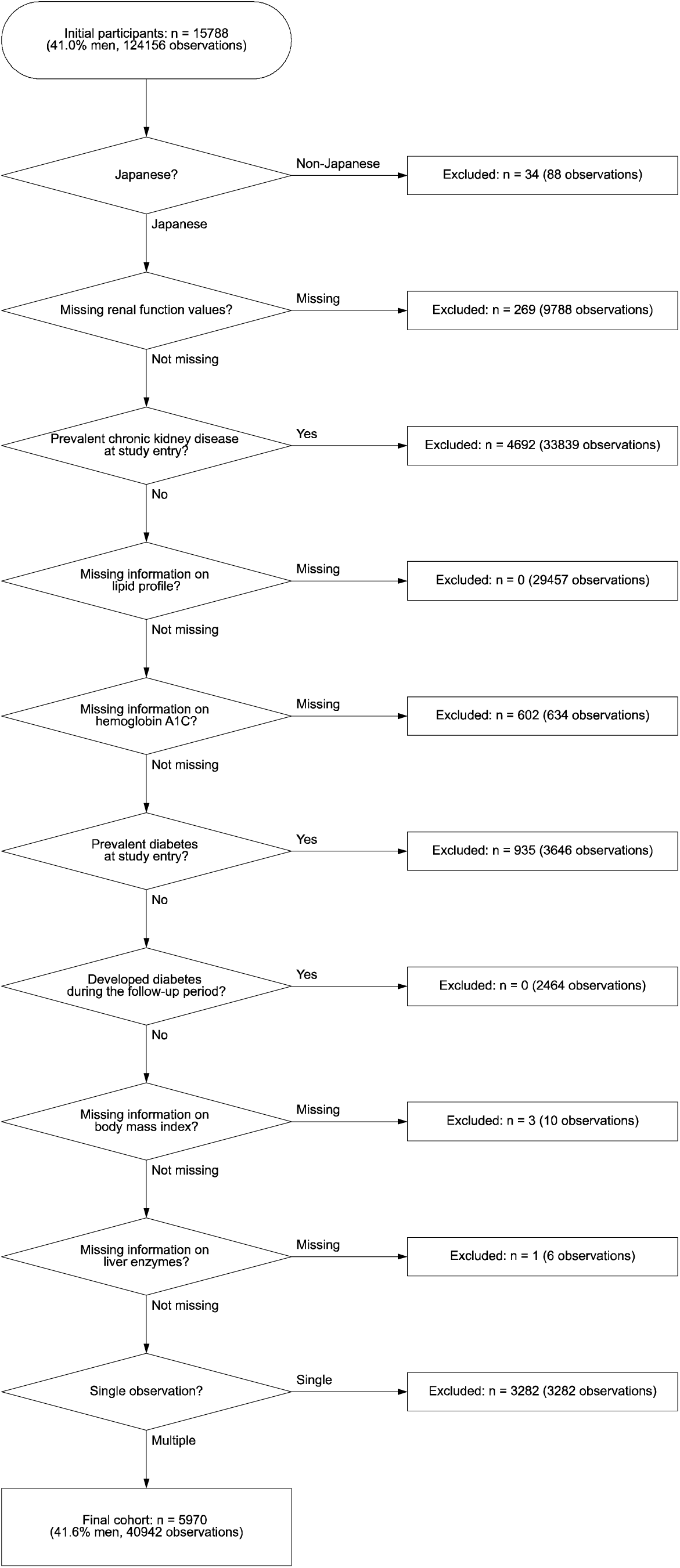
Flow diagram of the study participants.

**Figure 2.**
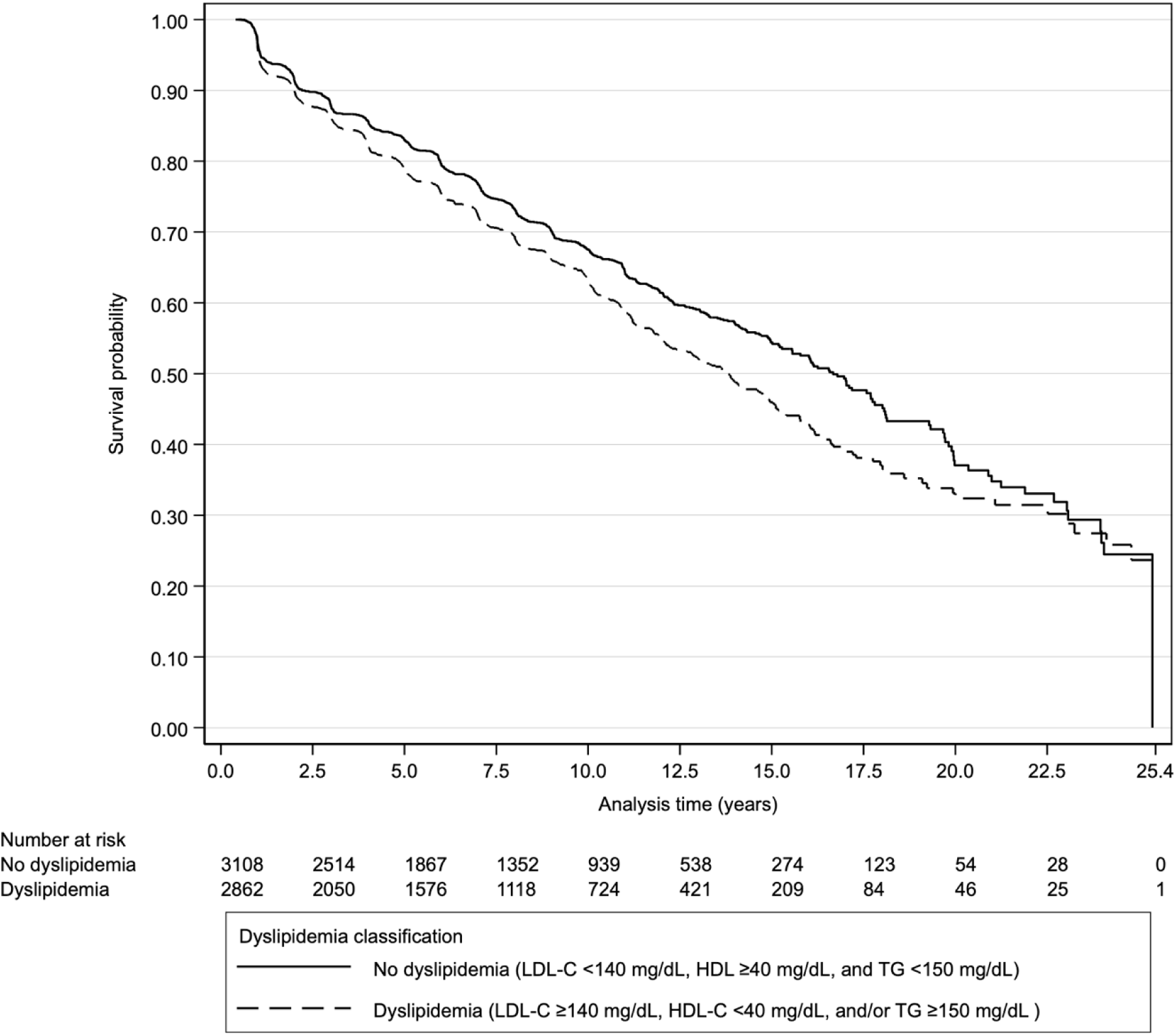
Kaplan–Meier curves stratified by dyslipidemia categories.

**Table 1** shows the distribution of variables by prevalent dyslipidemia during the follow-up. Prevalent dyslipidemia had a higher incidence rate than the normal lipid profile. The relationship between dyslipidemia and subsequent onset of CKD is shown in **Table 2** . Regardless of whether the covariates were adjusted, prevalent dyslipidemia showed a shorter survival time to later onset CKD than lipids in the normal range, and this survival time was 5% shorter (95% confidence interval [CI]: 3%–7%) in the full model.

**Table 1.** Descriptive statistics of all observations stratified by dyslipidemia classification of 5970 non-diabetic Japanese citizens of Zentsuji City (1998–2024)

|  | Dyslipidemia classification* |  |  |  |  |  |  |  |  |
| --- | --- | --- | --- | --- | --- | --- | --- | --- | --- |
|  | No dyslipidemia |  |  | Dyslipidemia |  |  | Total |  |  |
|  | Failure | PY | IR† | Failure | PY | IR† | Failure | PY | IR† |
| Time at risk |  |  |  |  |  |  |  |  |  |
| Total |  | 23077.1 |  |  | 19278.2 |  |  | 42355.3 |  |
| Minimum |  | 0.47 |  |  | 0.40 |  |  | 0.40 |  |
| Maximum |  | 25.4 |  |  | 24.7 |  |  | 25.5 |  |
| Mean |  | 5.39 |  |  | 4.85 |  |  | 7.09 |  |
| Median |  | 4.06 |  |  | 3.83 |  |  | 6.04 |  |
| Failure/IR† | 940 |  | 40.7 | 950 |  | 49.3 | 1890 |  | 44.6 |
| Sex |  |  |  |  |  |  |  |  |  |
| Female | 369 | 9253.9 | 39.9 | 400 | 7963.2 | 50.2 | 769 | 17217.0 | 44.7 |
| Male | 571 | 13823.2 | 41.3 | 550 | 11315.0 | 48.6 | 1121 | 25138.3 | 44.6 |
| Age, mean (SD) | 66.0 | (11.4) |  | 66.5 | (10.1) |  | 66.2 | (10.8) |  |
| BMI category‡ |  |  |  |  |  |  |  |  |  |
| Normal weight | 726 | 18969.6 | 38.3 | 642 | 14016.8 | 45.8 | 1368 | 32986.4 | 41.5 |
| Overweight or obesity | 214 | 4107.5 | 52.1 | 308 | 5261.4 | 58.5 | 522 | 9368.9 | 55.7 |
| Self-reported alcohol intake |  |  |  |  |  |  |  |  |  |
| Non- or seldom-drinker | 552.6 | 12316.4 | 44.9 | 591.2 | 11361.9 | 52.0 | 1143.8 | 23678.3 | 48.3 |
| Drinker | 387.4 | 10760.7 | 36.0 | 358.8 | 7916.3 | 45.3 | 746.2 | 18677.0 | 40.0 |
| Self-reported smoking status |  |  |  |  |  |  |  |  |  |
| Non- or ex-smoker | 853.8 | 19875.1 | 43.0 | 858.9 | 16623.7 | 51.7 | 1712.7 | 36498.8 | 46.9 |
| Smoker | 86.2 | 3202.0 | 26.9 | 91.1 | 2654.5 | 34.3 | 177.3 | 5856.5 | 30.3 |
| Blood pressure category¶ |  |  |  |  |  |  |  |  |  |
| Normal | 408 | 12577.1 | 32.4 | 387 | 8731.7 | 44.3 | 795 | 21308.8 | 37.3 |
| Hypertensive | 532 | 10500.1 | 50.7 | 563 | 10546.4 | 53.4 | 1095 | 21046.5 | 52.0 |
| HbA1c, mean (SD) | 5.53 | (0.34) |  | 5.58 | (0.33) |  | 5.55 | (0.34) |  |
| AST, mean (SD) | 23.3 | (17.2) |  | 23.9 | (9.92) |  | 23.6 | (14.4) |  |
| ALT, mean (SD) | 18.9 | (15.9) |  | 21.8 | (15.0) |  | 20.2 | (15.6) |  |
| GGT, mean (SD) | 29.1 | (34.4) |  | 34.4 | (46.3) |  | 31.5 | (40.2) |  |
| Residential district |  |  |  |  |  |  |  |  |  |
| East | 165.6 | 4332.8 | 38.2 | 187.9 | 3556.5 | 52.8 | 353.5 | 7889.3 | 44.8 |
| Tatsukawa | 162.5 | 3700.8 | 43.9 | 151.6 | 2996.1 | 50.6 | 314.1 | 6696.9 | 46.9 |
| South | 137.4 | 3898.9 | 35.2 | 156.7 | 3376.7 | 46.4 | 294.1 | 7275.6 | 40.4 |
| Fudeoka | 101.0 | 2916.7 | 34.6 | 96.4 | 2274.3 | 42.4 | 197.4 | 5191.1 | 38.0 |
| Central | 103.5 | 2292.9 | 45.2 | 117.6 | 2096.2 | 56.1 | 221.1 | 4389.1 | 50.4 |
| West | 97.7 | 2351.0 | 41.6 | 89.8 | 1775.0 | 50.6 | 187.6 | 4126.0 | 45.5 |
| Yoshiwara | 88.9 | 2075.5 | 42.8 | 85.0 | 1828.6 | 46.5 | 173.9 | 3904.1 | 44.5 |
| Yogita | 83.5 | 1508.6 | 55.3 | 64.8 | 1374.6 | 47.1 | 148.3 | 2883.2 | 51.4 |

Abbreviations: ALT, alanine aminotransferase; AST, aspartate aminotransferase; BMI, body mass index; DBP, diastolic blood pressure; GGT, gamma-glutamyl transferase; HbA1c, hemoglobin A1C; HDL, high-density lipoprotein cholesterol; IR, incidence rate; LDL, low-density lipoprotein cholesterol; SBP, systolic blood pressure; SD, standard deviation; TG, triglycerides.
Results for multiple imputed variables (self-reported alcohol intake, self-reported smoking status, and residential district) are averaged over 30 imputations.
\*Dyslipidemia is defined as LDL $\geq 140$ mg/dL, HDL $< 40$ mg/dL, and/or TG $\geq 150$ mg/dL.
†Incidence rate is reported per 1000 person-years.
‡Overweight or obesity is defined as a BMI $\geq 25$ kg/m<sup>2</sup>.
¶Hypertension is defined as SBP $\geq 130$ mmHg and/or DBP $\geq 80$ mmHg.

**Table 2.** Association between dyslipidemia and subsequent development of chronic kidney disease (eGFR < 60 mL/min/1.73 m ), using the Weibull accelerated failure-time model (Zentsuji, Kagawa Prefecture, Japan, 1998–2024)

| Variable | Total (n=5970) |  | IR¶ | Crude<br>TR (95% CI) | Model 1<br>aTR (95% CI) | Model 2<br>aTR (95% CI) | Model 3<br>aTR (95% CI) | Model 4<br>aTR (95% CI) |
| --- | --- | --- | --- | --- | --- | --- | --- | --- |
|  | Person-years | Failures |  |  |  |  |  |  |
| No dyslipidemia (reference) | 23077.1 | 940 | 40.7 | 1.00 | 1.00 | 1.00 | 1.00 | 1.00 |
| Dyslipidemia† | 19278.2 | 950 | 49.3 | 0.97 (0.95–0.98) | 0.96 (0.95–0.98) | 0.97 (0.95–0.98) | 0.96 (0.95–0.98) | 0.95 (0.93–0.97) |

Abbreviations: ALT, alanine aminotransferase; AST, aspartate aminotransferase; aTR, adjusted time ratio; BMI, body mass index; CI, confidence interval; DBP, diastolic blood pressure; eGFR, estimated glomerular filtration rate; GGT, gamma-glutamyl transferase; HbA1c, hemoglobin A1C; HDL-C, high-density lipoprotein cholesterol; IR, incidence rate; LDL-C, low-density lipoprotein cholesterol; SBP, systolic blood pressure; TG, triglycerides; TR, time ratio.
\*Dyslipidemia was defined as LDL-C ≥140 mg/dL, HDL-C < 40 mg/dL, and/or TG ≥150 mg/dL.
†Overweight or obesity was defined as BMI ≥25 kg/m<sup>2</sup>.
‡Hypertension was defined as SBP ≥130 mmHg and/or DBP ≥80 mmHg.
¶Incidence rate is reported per 1000 person-years.
Model 1 was adjusted for sex (male[reference]/female) and age. A multiplicative term (dyslipidemia\* × sex) was added.
Model 2 was adjusted for both variables of Model 1, overweight or obesity (no[reference]/yes)†, self-reported smoking status (non- or ex-smoker[reference]/smoker), and self-reported alcohol intake (non- or seldom-drinker[reference]/drinker). A multiplicative term (dyslipidemia\* × sex) was added.
Model 3 was adjusted for all variables of Model 2, hypertension (no[reference]/yes)‡, HbA1c, AST, ALT, and GGT. A multiplicative term (dyslipidemia\* × sex) was added.
Model 4 was adjusted for all variables of Model 3 and the residential area of the city. The multiplicative terms (dyslipidemia\* × sex, dyslipidemia\* × hypertension‡) were added.

### Sensitivity analyses

When we separately evaluated the associations between LDL-C, HDL-C, and TG concentrations and the development of CKD in the first sensitivity analysis (**Table S1** ), HDL-C concentrations <40 mg/dL and TG concentrations ≥150 mg/dL had a 5% shorter survival time (95% CI: 3%–7%) to CKD onset than the normal lipid profile (HDL-C concentrations ≥60 mg/dL and TG concentrations <100 mg/dL). In contrast, elevated LDL-C concentration categories (120–<140 mg/dL and ≥140 mg/dL) had an adjusted time ratio of 1.00 (95% CI: 0.99–1.02), which was almost the null value, compared with LDL-C concentrations <120 mg/dL.

**Table S2** shows the results of the second sensitivity analysis that examined how the results changed after using the definition of CKD of an eGFR <60 mL/min/1.73 m^2^ and/or prevalent proteinuria ≥1+. The results were slightly attenuated compared with those of the main results shown in **Table 2** . Prevalent dyslipidemia shortened the survival time to CKD onset by 4% (95% CI: 2%–6%) in the full model.

## Discussion

This 26-year longitudinal study showed that prevalent dyslipidemia reduced the survival time to CKD onset by 5%, even in a non-diabetic population. This study adds further evidence that dyslipidemia may be a risk factor for new-onset CKD late in life, even in a non-diabetic Asian population. The strengths of this study are that it included a mono-ethic Asian adult population without diabetes, had a long follow-up time of up to approximately 26 years (**Table 1** ), and used the parametric accelerated failure-time model to attempt to reduce the effect of changes in covariates over time during the follow-up.

A previous UK Biobank cohort study showed an association between lipid concentrations (LDL-C, HDL-C, and TG) and the risk of CKD[4]. Similarly, a study in the general Chinese population showed an association between dyslipidemia and CKD development, with higher TG concentrations increasing the risk of CKD[3]. However, the results of LDL-C concentrations in these studies are different from those in our study. Our findings regarding the associations between HDL-C (<40 mg/dL) and TG (≥150 mg/dL) concentrations and a shorter survival time to CKD onset are in agreement with these two previous studies, but elevated LDL-C concentrations were not related to survival (**Table S1** ).

There are four possible reasons for the inconsistent LDL-C results between studies. First, only a few studies examined the relationship between dyslipidemia and the later development of CKD, and all of these studies used different definitions of dyslipidemia[3,4]. This study used the cutoff values in accordance with the Japan Atherosclerosis Society guideline, the UK Biobank study standardized lipid profiles with a 1-SD increase, and the Chinese study used quartiles as cutoff values for the lipid profile[3,4,19]. This difference in the standard may be part of the reason why the LDL-C results are different between studies.

Second, differences in the study participants may have affected the results. This study only included non-diabetic Japanese participants, and the UK Biobank study and Chinese general population study included participants with and without diabetes[3,4]. Furthermore, the current study and the Chinese study included East Asian populations, but the UK Biobank study only included White participants. Previous studies reported that the prevalence of diabetes was higher in East Asians than in non-Hispanic Whites[16,33]. Diabetes often coexists with dyslipidemia and is the leading cause of CKD[16]. Therefore, differences in characteristics of the target populations (i.e., race and whether diabetes was included) may have affected the LDL-C results, which may have been overestimated by residual confounding of diabetes as in previous studies[3,4,20].

Third, differences in the follow-up time between the studies may have affected the LDL-C results. The follow-up period (median follow-up) was 1998–2024 (6.04 years) in this study, 2006–2019 (10.7 years) in the UK Biobank study, and 2011–2016 (the median follow-up time was not shown) in the Chinese study[3,4]. A longer follow-up time may induce built-in selection bias, leading to underestimating results[34]. Therefore, the results of this study and the UK Biobank study may have been underestimated.

Fourth, differences in selecting variables for adjustments may have affected the LDL-C results. Previous studies further adjusted for variables that this study did not include, such as statin use, uric acid concentrations, and the fasting time[3,4]. Statins, which are cholesterol-lowering medications, reduce LDL-C concentrations, which also attenuate the risk of a decline in kidney function[35]. In this situation, more participants with dyslipidemia would have been classified as normolipidemic and more participants with CKD would have been classified as non-CKD. This classification would have led to differential misclassification and an overestimated association. Additionally, higher blood uric acid concentrations are associated with elevated lipid profiles and a decline in renal function. Therefore, the association between dyslipidemia and incident CKD may have been overestimated because we did not adjust for the variables’ effects[36,37]. Furthermore, this study included fasting and non-fasting participants. All associations in this study may have been biased toward the null value because non-fasting participants tend to be misclassified in the dyslipidemia group[38]. The effects of unavailable information, as indicated above, may cancel each other out. Therefore, the misclassification in this study is assumed to be non-differential. However, there may have been residual effects from other variables, which may explain the inconsistent results of LDL-C in this study and in previous studies[3,4].

There are several limitations to this study. First, the characteristics of the participants in this study and in the previous two studies mentioned above were different, resulting in the use of different definitions of dyslipidemia[3,4]. The participants in our study were older than those in the previous two studies, which may have led to overestimated associations in our study[20]. To mitigate this issue, we adjusted for age in all analyses. This study included only non-diabetic Japanese people, while the previous two studies included non-Hispanic Whites and Chinese with and without diabetes. Various outcomes (e.g., diabetic nephropathy, stroke, heart failure, hospitalization, and death) related to prevalent diabetes and race affect the development of dyslipidemia and CKD[8,10,18,20,39]. Furthermore, the participation in checkups in this study was completely voluntary. Consequently, people with illnesses serious enough to prevent them from performing checkups and the effects of diabetes complications were absent[40]. Therefore, all results of this study are considered to have been underestimated, and generalizing the results to other populations with different races and age structures is limited. However, our findings may be applicable to developed countries with non-diabetic older Asian populations.

Second, the vulnerability of the eGFR may have affected our results. Therefore, we used the CKD criteria using an eGFR <60 mL/min/1.73 m^2^ and proteinuria ≥1+ in the second sensitivity analysis (**Table S2** )[29]. These results were consistent with the main analysis (**Table 2**) , which indicated that these differences in the definition of CKD did not affect the results. However, there are other useful indicators of CKD such as albuminuria, which were unavailable in this study[20]. Albuminuria (e.g., the urinary albumin-to-creatinine ratio) indicates elevated protein and is recommended for screening for CKD in addition to using the eGFR[41]. A previous study in the Japanese general population (n=22,975) showed that the prevalence of CKD was higher when using the eGFR and urinary albumin-to-creatinine ratio than when using the eGFR and proteinuria[42]. Therefore, missing values for the urinary albumin-to-creatinine ratio can cause misclassification of CKD as non-CKD, which may have resulted in an underestimation of associations in this study.

Finally, we were unable to adjust for unmeasured important covariates, such as medications, the prevalence of diseases that affect both exposure and outcome variables, physical inactivity, dietary patterns, hospitalization, and death[20,43,44]. This is a limitation that cannot be ignored, but these covariates cannot be adjusted for. In particular, hospitalization and death are competing risks that preclude obtaining incident CKD data. Therefore, if competing risks were present in this study, the results could have been underestimated.

## Conclusions

This longitudinal study shows that dyslipidemia is an independent risk factor for the later development of CKD in a non-diabetic Japanese population, with a 5% shorter survival time to the development of CKD. Elevated TG concentrations ≥150 mg/dL and low HDL-C concentrations <40mg/dL, but not elevated LDL-C concentrations ≥140 mg/dL, also result in a 5%–6% shorter survival to CKD than the reference category. Our findings suggest that controlling the lipid profile to the normal range might be beneficial for future public health efforts to prevent CKD.

## Data Availability

All data generated or analyzed during this study are included in this published article and its Supplementary files.

## Ethic

The data was anonymized before receipt. The Ethics Committee of Okayama University Graduate School of Medicine, Dentistry and Pharmaceutical Sciences and Okayama University Hospital approved this study (No. K1708-040). The Ethics committee waived the need for informed consent. This research followed the Declaration of Helsinki and Japanese Ethical Guidelines for Medical and Biological Research involving Human Subjects.

## Acknowledgments

We thank all participants in this study, Ayaka Nakatsu, Masako Matsumoto, Mayumi Kitadani, and the local government officers of Zentsuji City who kindly supported our research. We thank Ellen Knapp, PhD, from Edanz (https://jp.edanz.com/ac) for editing a draft of this manuscript.

## Author Contributions

Conceptualization, Y.O.; methodology, Y.O., T.M.; formal analysis, Y.O.; investigation, Y.O.; data curation, Y.O.; writing—original draft preparation, Y.O.; writing—review and editing, Y.O., T.M.; visualization, Y.O.; supervision, T.M.; project administration, Y.O. This manuscript has been read and approved by both authors.

