## Supplementary Tables for "Dyslipidemia and development of chronic kidney disease in non-diabetic Japanese adults: a 26-year, community-based, longitudinal study"

**Table S1**. Associations between LDL-C, HDL-C, and TG concentrations and subsequent development of chronic kidney disease (eGFR <60 mL/min/1.73 $m^2$), using the Weibull accelerated failure-time model (Zentsuji, Kagawa Prefecture, Japan, 1998–2024)

| | Total (n=5970) | | | Crude | Model 1 | Model 2 | Model 3 | Model 4 |
|---|---|---|---|---|---|---|---|---|
| Variables | Person-years | Failures | IR‡ | TR (95% CI) | aTR (95% CI) | aTR (95% CI) | aTR (95% CI) | aTR (95% CI) |
| **LDL-C**[a] | | | | | | | | |
| <120 mg/dL (reference) | 18163.8 | 837 | 46.1 | 1.00 | 1.00 | 1.00 | 1.00 | 1.00 |
| 120–<140 mg/dL | 11218.6 | 494 | 44.0 | 1.00 (0.98–1.02) | 1.00 (0.99–1.02) | 1.01 (1.00–1.02) | 1.01 (1.00–1.02) | 1.01 (1.00–1.02) |
| ≥140 mg/dL | 12972.8 | 559 | 43.1 | 1.00 (0.98–1.02) | 0.98 (0.97–1.00) | 1.00 (0.99–1.02) | 1.00 (0.99–1.02) | 1.00 (0.99–1.02) |
| **HDL-C**[b] | | | | | | | | |
| <40 mg/dL | 2376.4 | 168 | 70.7 | 0.93 (0.90–0.96) | 0.95 (0.93–0.97) | 0.95 (0.93–0.97) | 0.95 (0.93–0.97) | 0.95 (0.93–0.97) |
| 40–<60 mg/dL | 18077.5 | 875 | 48.4 | 0.98 (0.96–0.99) | 0.98 (0.97–0.99) | 0.99 (0.97–1.00) | 0.98 (0.97–1.00) | 0.98 (0.97–1.00) |
| ≥60 mg/dL (reference) | 21901.5 | 847 | 38.7 | 1.00 | 1.00 | 1.00 | 1.00 | 1.00 |
| **TG**[c] | | | | | | | | |
| <100 mg/dL (reference) | 21257.9 | 804 | 37.8 | 1.00 | 1.00 | 1.00 | 1.00 | 1.00 |
| 100–<150 mg/dL | 12692.3 | 627 | 49.4 | 0.97 (0.95–0.99) | 0.98 (0.97–0.99) | 0.98 (0.97–0.99) | 0.98 (0.97–1.00) | 0.98 (0.96–1.00) |
| ≥150 mg/dL | 8405.1 | 459 | 54.6 | 0.94 (0.92–0.96) | 0.96 (0.95–0.98) | 0.97 (0.96–0.98) | 0.95 (0.93–0.97) | 0.95 (0.93–0.97) |

Abbreviations: ALT, alanine aminotransferase; AST, aspartate aminotransferase; aTR, adjusted time ratio; BMI, body mass index; CI, confidence interval; DBP, diastolic blood pressure; eGFR, estimated glomerular filtration rate; GGT, gamma-glutamyl transferase; HbA1c, hemoglobin A1C; HDL-C, high-density lipoprotein cholesterol; IR, incidence rate; LDL-C, low-density lipoprotein cholesterol; SBP, systolic blood pressure; TG, triglycerides; TR, time ratio.

* Overweight or obesity was defined as BMI ≥25 kg/$m^2$.

† Hypertension was defined as SBP ≥130 mmHg and/or DBP ≥80 mmHg.

‡ Incidence rate is reported per 1000 person-years.

a Model 1 was adjusted for sex (male[reference]/female) and age. A multiplicative term (LDL-C × sex) was added.
Model 2 was adjusted for both variables of Model 1, overweight or obesity (no[reference]/yes)*, self-reported alcohol intake (non- or seldom-drinker[reference]/drinker), and self-reported smoking status (non- or ex-smoker[reference]/smoker).
Model 3 was adjusted for all variables of Model 2, hypertension (no[reference]/yes)†, HbA1c, AST, ALT, and GGT.
Model 4 was adjusted for all variables of Model 3 and the residential area of the city.

b Model 1 was adjusted for sex (male[reference]/female) and age.
Model 2 was adjusted for both variables of Model 1, overweight or obesity (no[reference]/yes)*, self-reported alcohol intake (non- or seldom-drinker[reference]/drinker), and self-reported smoking status (non- or ex-smoker[reference]/smoker).
Model 3 was adjusted for all variables of Model 2, hypertension (no[reference]/yes)†, HbA1c, AST, ALT, and GGT.
Model 4 was adjusted for all variables of Model 3 and the residential area of the city.

c Model 1 was adjusted for sex (male[reference]/female) and age.
Model 2 was adjusted for both variables of Model 1, overweight or obesity (no[reference]/yes)*, self-reported alcohol intake (non- or seldom-drinker[reference]/drinker), and self-reported smoking status (non- or ex-smoker[reference]/smoker).
Model 3 was adjusted for all variables of Model 2, hypertension (no[reference]/yes)†, HbA1c, AST, ALT, and GGT. A multiplicative term (TG × hypertension†) was added.
Model 4 was adjusted for all variables of Model 3 and the residential area of the city. A multiplicative term (TG × hypertension†) was added.

**Table S2**. Associations between dyslipidemia and subsequent development of chronic kidney disease (eGFR <60 mL/min/1.73 $m^2$ and/or proteinuria ≥1+), using the Weibull accelerated failure-time model (Zentsuji, Kagawa Prefecture, Japan, 1998–2024)

| | Total (n=5758) | | | Crude | Model 1 | Model 2 | Model 3 | Model 4 |
|---|---|---|---|---|---|---|---|---|
| Variable | Person-years | Failures | IR¶ | TR (95% CI) | aTR (95% CI) | aTR (95% CI) | aTR (95% CI) | aTR (95% CI) |
| No dyslipidemia (reference) | 21389.0 | 1098 | 51.3 | 1.00 | 1.00 | 1.00 | 1.00 | 1.00 |
| Dyslipidemia* | 17777.9 | 1103 | 62.0 | 0.96 (0.95–0.98) | 0.96 (0.94–0.97) | 0.96 (0.94–0.98) | 0.96 (0.94–0.98) | 0.96 (0.94–0.98) |

Abbreviations: ALT, alanine aminotransferase; AST, aspartate aminotransferase; aTR, adjusted time ratio; BMI, body mass index; CI, confidence interval; DBP, diastolic blood pressure; eGFR, estimated glomerular filtration rate; GGT, gamma-glutamyl transferase; HbA1c, hemoglobin A1C; HDL-C, high-density lipoprotein cholesterol; IR, incidence rate; LDL-C, low-density lipoprotein cholesterol; SBP, systolic blood pressure; TG, triglycerides; TR, time ratio.

*Dyslipidemia was defined as LDL-C ≥140 mg/dL, HDL-C < 40 mg/dL, and/or TG ≥150 mg/dL.

†Overweight or obesity was defined as BMI ≥25 kg/$m^2$.

‡Hypertension was defined as SBP ≥130 mmHg and/or DBP ≥80 mmHg.

¶Incidence rate is reported per 1000 person-years.

Model 1 was adjusted for sex (male[reference]/female) and age. A multiplicative term (dyslipidemia* × sex) was added.

Model 2 was adjusted for both variables of Model 1, overweight or obesity (no[reference]/yes)†, self-reported alcohol intake (non- or seldom-drinker[reference]/drinker), and self-reported smoking status (non- or ex-smoker[reference]/smoker). A multiplicative term (dyslipidemia* × sex) was added.

Model 3 was adjusted for all variables of Model 2, hypertension (no[reference]/yes)‡, HbA1c, AST, ALT, and GGT. A multiplicative term (dyslipidemia* × sex) was added.

Model 4 was adjusted for all variables of Model 3 and the residential area of the city. A multiplicative term (dyslipidemia* × sex) was added.
